# Altered cerebrovascular response to breath holding in thoracolumbar spinal cord injury measured using functional near-infrared spectroscopy

**DOI:** 10.64898/2026.03.12.26348285

**Authors:** Keerthana Deepti Karunakaran, Donna Y Chen, Nancy D. Chiaravalloti, Bharat B. Biswal

## Abstract

Spinal cord injury (SCI) is associated with cardiovascular deficits that affect cerebral blood flow, cerebral perfusion, and cerebrovascular control. While several studies use neuroimaging techniques such as functional magnetic resonance imaging (fMRI) to understand neuroplasticity following SCI, more work needs to be done to evaluate the cerebrovascular changes following SCI. Understanding these effects using neuroimaging is essential as these deficits also affect neurovascular coupling and how we interpret neuroplasticity measured based on neuroimaging. Hence, we conducted a pilot study in twelve healthy males and thirteen males with thoracolumbar SCI using functional near-infrared spectroscopy (fNIRS) to understand the effects of breath-holding induced hypercapnia on the hemodynamics of the sensorimotor cortex and prefrontal cortex (PFC) after SCI. Participants performed 30 seconds of regular breathing alternated by 15 seconds of breath-holding for 5 minutes. Compared to controls, the SCI group presented with a greater initial decrease in oxy-hemoglobin concentration change and a delayed subsequent increase in oxy-hemoglobin concentration change in response to hypercapnia at p<. Additionally, the net increase in oxy-hemoglobin concentration change following BH in the PFC was negatively correlated with the level of injury at p=0.005, where higher levels of injury were associated with a smaller increase in oxy-hemoglobin concentration following hypercapnia. These findings confirm that a) SCI, including lower levels of injury (below T6) are associated with cerebrovascular changes that are quantifiable using fNIRS, and b) fNIRS could be a robust tool to understand the neuroplastic and cerebrovascular changes in people with SCI.

## Introduction

Spinal Cord Injury (SCI) disrupts supraspinal sympathetic control of the heart, causing an upregulation of the parasympathetic control through the intact efferent vagal nerve^1,2^. Hence, SCI is associated with cardiovascular deficits such as low resting blood pressure, orthostatic hypotension, cardiac arrhythmia, and autonomic dysreflexia^3^. These deficits vary in severity depending on the level and completeness of injury, where injuries above the splanchnic sympathetic outflow level (T5-T6) result in the loss of sympathetic preganglionic neurons to the heart and, therefore, experience more severe deficits^4^. Consequently, this imbalance in the autonomic system is associated with disruption of cerebrovascular control and increased risk of stroke and ischemic damage in these individuals^5^. Lack of mobility, functional degeneration, and arterial stiffening during chronic stages of injury is believed to further contribute to cardiovascular complications independent of or secondary to autonomic deficits. Dong-II Kim and group proposed that SCI is characterized by abnormal perfusion in response to changes in arterial pressure, arterial gas, and metabolic demand compared to individuals without injury, collectively known as cerebrovascular control^6^. It is suggested that dysregulation in cardiovascular and cerebrovascular control may also influence neurocognition which may directly or indirectly affect their recovery and personal burden ^7,8^. Existing literature on the SCI population is consistent and indicates that dynamic cerebrovascular regulation is impaired after SCI, which is the rapid regulation of cerebral blood flow in response to changes in blood pressure, blood CO_2_ levels and neuronal activity (neurovascular coupling)^9,10^. However, current methods of cerebral blood flow assessments are either impractical for naturalistic settings (e.g., fMRI)^11–13^, or lack the spatial resolution (e.g., transcranial doppler)^14^ to perform a comprehensive evaluation of cerebral blood flow during functional tasks. In the current study, we investigated the cerebral hemodynamic response to breath-hold-induced hypercapnia using a portable optical brain imaging—functional near-infrared spectroscopy (fNIRS) in 13 males with chronic thoracolumbar SCI compared to 13 age-matched healthy males. fNIRS technology, based on the principle of neurovascular coupling, offers a portable, economical, and flexible approach to quantifying the cerebrovascular response in both in-lab and out-of-lab settings^15,16^. We hypothesized that the hemodynamic response to breath holding measured using fNIRS is sensitive to the alterations in cerebrovascular reactivity after SCI and could offer critical insight into the extent of these impairments in lower-level injuries and chronic stages of the disease.

## Methods

Participants comprised thirteen males with thoracolumbar spinal cord injury (48.4 years) and 12 age-matched healthy males (47.6 years). All individuals underwent a 5-minute breath-holding paradigm where they performed 30 seconds of normal breathing, alternated with 15 seconds of breath-holding for a total duration of 300 seconds. A 32-channel fNIRS system with 690 and 830 nm lasers (CW6, Tech En Inc.) was used at a sampling frequency of 50Hz. Oxy-, deoxy-, and total-hemoglobin concentration changes from the prefrontal cortex, pre-motor, and sensorimotor cortices were recorded. The 32 channels were divided into seven regions of interest (ROI) covering prefrontal cortex (PFC), supplementary motor area (SMA), medial sensorimotor region (SMN), mediolateral SMN and lateral SMN. The fNIRS data was preprocessed using in-house scripts on the MATLAB R2022b platform. The details of the participants, inclusion/exclusion criteria, Optode setup, regions of interest (ROI), and data preprocessing methodology are described elsewhere^17^. The study staff explained and obtained written informed consent from each participant based on the guidelines outlined by the institutional review board of the New Jersey Institute of Technology. A block averaging technique was utilized to generate the hemodynamic response in each ROI and each subject. A block was defined as the 5 seconds before the start of the breath holding, the 15 seconds of breath holding, and the 30 seconds of rest following stimuli. The maximum decrease in the hemodynamic response during breath holding (5 to 15 seconds after onset of task) and maximum increase in the hemodynamic response after breathing resumed (at 15 seconds after onset of breath holding) were calculated as parameters of cerebrovascular reactivity for each channel in each subject. Further, to identify changes in the latency of breath hold activation, the time of maximum increase in ΔHbO concentration was estimated. An independent sample t-test was performed to identify differences in the breath-holding based CVR measures (maximum increase in ΔHbO, maximum decrease in ΔHbO, time to maximum increase in ΔHbO) between HC and SCI groups. A one-way ANOVA was performed to understand the effects of duration since injury (continuous measure), incidence of neuropathic pain (categorical measure), and level of injury (number of affected spinal segments as a continuous measure) on the CVR measures. A statistical threshold of alpha= 0.05 with a Benjamini Hochberg False Discovery Rate-based multiple comparison correction was implemented^18^.

## Results

The average hemodynamic response to 15 seconds of breath-holding in the 7 ROIs is shown in **Figure 1**. BH induced hypercapnia in healthy volunteers was generally associated with an initial decrease in ΔHbO concentration followed by a delayed increase in ΔHbO concentration. This initial decrease in ΔHbO concentration during BH was more pronounced in SCI participants than in HC participants (see a more significant reduction in ΔHbO time series at 0 to 15 secs). Further comparison of the characteristics of the HDR between the two groups using independent sample t-tests confirmed a statistically significant maximum decrease in ΔHbO concentration during BH in left sensorimotor regions at FDR-p<0.05 (uncorr-p=0.008) and in the supplementary motor area and PFC at p<0.05 (**Figure 2**). Though the maximum increase in the ΔHbO time series following BH was comparable between the two groups, the time to attain a maximum increase in ΔHbO time series was longer in the SCI group (See **Figure 2C**). The net increase in ΔHbO concentration was also negatively correlated with the level of injury at FDR-p<0.05 (uncorr-p=0.005), where higher levels of injury were associated with lower levels of ΔHbO concentration following BH (**Figure 2B**).

**Figure 1.**
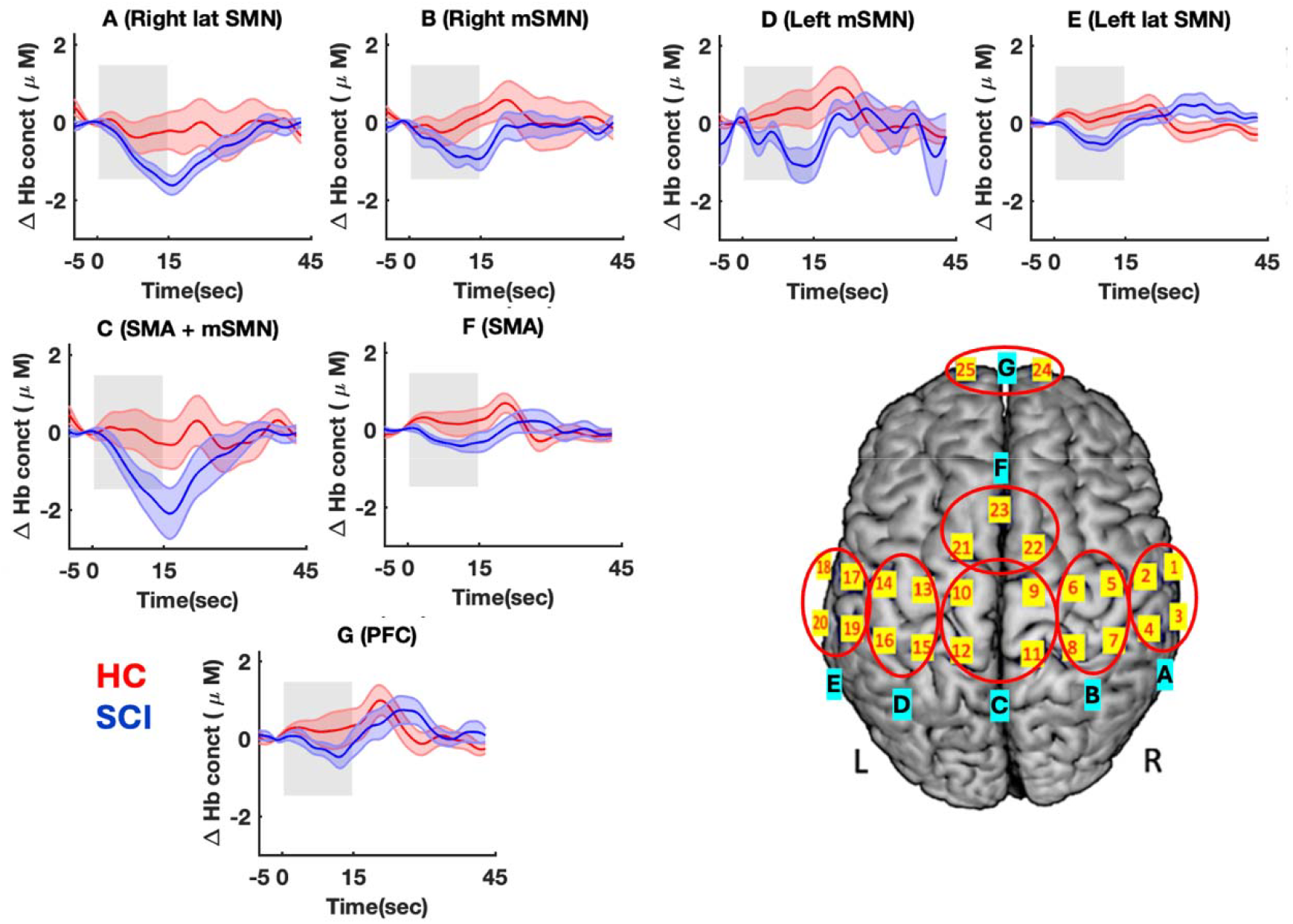
Average hemodynamic response (ΔHbO concentration) to 15 second breath-holding in seven regions of interest are shown for SCI patients (n=13) and healthy volunteers (n=12). The channels within each region of interest (A to G) are indicated by a red circle on the brain map. The shaded grey area on the hemodynamic plots represents the duration of breath-holding.

**Figure 2.**
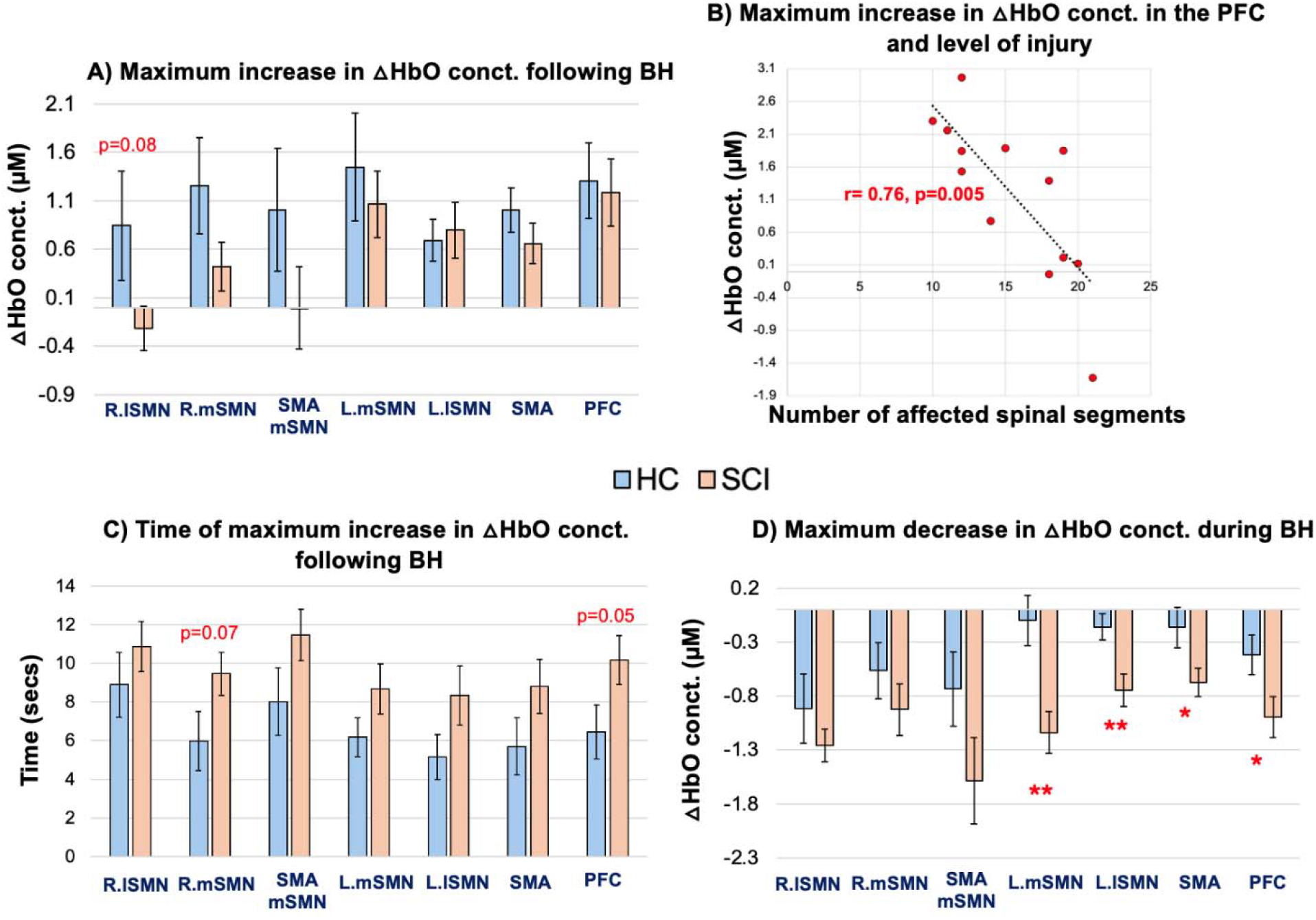
Metrics of cerebrovascular reactivity quantified from the hemodynamic response (ΔHbO concentration) to 15 second breath-holding in seven regions of interest are shown for SCI patients (n=13) and healthy volunteers (n=12). (A) Maximum increase in ΔHbO concentration, (B) Relationship between maximum increase in ΔHbO concentration in the PFC region and the level of injury (calculated as number of affected spinal segments) at FDR-p<0.05 (r=0.76, p=0.005), (C) Time of maximum increase in ΔHbO concentration following BH, and (D) Maximum decrease in ΔHbO concentration during BH. * indicates p<0.05, and ** indicates FDR-p<0.05 (p=0.008).

## Discussion

Breath-holding is a reliable and well-established vasoactive stimulus to induce hypercapnia naturally^19^. Neuroimaging studies frequently implement it to investigate cerebrovascular reactivity^20–22^. Our study using fNIRS shows that thoracolumbar SCI is associated with reduced cerebrovascular reactivity, measured in the form of a more significant initial decrease in oxyhemoglobin concentration change and a delayed subsequent increase in oxy-hemoglobin concentration following BH. This is consistent with an MRI study using controlled CO_2_ inhalation^23^. An impaired breath-hold response was anticipated in the SCI group due to the sympathetic imbalance below the level of injury and reduced venous return from the blood vessels in the paralyzed/affected body, leading to overall instability in maintaining arterial blood pressure^6^. This imbalance in the sympathetic and parasympathetic outflow is thought to result in reduced resting blood pressure, orthostatic hypotension, and autonomic dysreflexia. Mainly, injuries above the T5 spinal level are prone to more severe instability of cerebral perfusion due to the direct innervation of cerebral vessels by sympathetic nerve fibers arising from these spinal segments (T1-T5 synapsing via cervical ganglia)^6^. Interestingly, similar peak HbO responses in both SCI and HC groups suggest that the overall cerebral blood flow i.e., autoregulatory action in response to hypercapnia is likely intact. However, the reduced reactivity or sensitivity to arterial CO_2_ resulting from breath holding, seen as a greater initial decrease in HbO, suggests the time to attain autoregulation is affected. This delay in autoregulation was observed at all levels of the thoracolumbar injury, including those below the T6 level. These findings are consistent with previous studies that show an intact static cerebral autoregulation but an impaired dynamic cerebral autoregulation^9,24,25^. Static autoregulation is the overall change in cerebral perfusion in response to blood pressure (or metabolic/CO_2_ changes), whereas the dynamic autoregulation reflects the rapid regulation and latency of the cerebral perfusion change. Impairment in cerebrovascular autoregulation is thought to first affect the latency (dynamic) of the response before affecting the overall efficiency of the autoregulation. Furthermore, the peak HbO response in our data was negatively associated with the level of injury where higher levels of injury were associated with lower levels of HbO change following hypercapnia. Thus, it is possible that higher levels of injury will exhibit greater deficits in dynamic autoregulation that may extend to static autoregulation after prolonged durations of injury. Similar effects are also reported after traumatic brain injury^26^.

### Applications of fNIRS in SCI population

fNIRS is an easy-to-use, portable, and cost-effective brain imaging tool that allows reliable continuous monitoring of changes in cortical hemodynamic activity^15,27^. fNIRS, like fMRI, indirectly measures brain function through the principle of neurovascular coupling, where neuronal activity results in metabolic consumption of oxygenated blood that is replenished by an influx of blood flow and increase in blood volume^27^. Simultaneous fNIRS-MRI studies have shown good agreement between the two modalities, suggesting fNIRS could be a good alternative to fMRI for interventions that require repeated, long-durations of assessments in naturalistic or out-of-lab settings^28–30^. Indeed, fMRI has better spatial coverage and resolution than fNIRS. However, fNIRS offers a unique opportunity for clinical applications, where continuous, repeated, outpatient/out-of-lab brain assessments are feasible with a moderate spatial resolution and high temporal resolution. Several studies report a link between hypotension, cerebrovascular dysregulation, and cognitive impairment in people with SCI ^8,31,32^. fNIRS recordings offer robust measures of prefrontal cortex with excellent reproducibility and could be used to understand the neural substrates of cognitive-emotional impairments after SCI^32^. fNIRS could be particularly suited for SCI population due to its flexibility in recording brain measures during supine, sitting or ambulatory tasks. Considering majority of the rehabilitation treatments involve ambulatory tasks on treadmill or similar setup, fNIRS could be used to understand the neurobiology of therapeutic techniques as well as the efficiency of the various approaches^33,34^. In addition, brain computer interfaces deriving from fNIRS could be used as a communication device or to control neuroprosthetics for people with SCI ^35,36^. The technique may also be combined with other modalities, such as electroencephalography (EEG) ^37–39^, transcranial magnetic stimulation^40^, biophysical^41^, and biomechanical tools, in various environments, including immersive virtual reality^42^ to evaluate integrative therapy^43,44^ as well as investigate the less-understood aspects of SCI such as abnormal emotional processing^45^, neuropathic pain^46^, phantom limb syndrome^47^ etc.

In conclusion, the benefits of fNIRS are numerous and well-established. This study focusing on a small sample of males with thoracolumbar SCI establishes the utility of fNIRS in investigating cerebrovascular dysregulation in SCI. Further studies on a larger subject pool including cervical injury are needed to confirm our findings, and to understand its implications for cognitive impairment, autonomic imbalance, and cardiovascular events such as stroke.

## Data Availability

All data produced in the present study are available upon reasonable request to the authors.

## Data Availability

Data will be made available upon request to the corresponding author.

## Acknowledgments

Portions of this article are published as part of the doctoral dissertation of KDK.

## Authors’ Contributions

The authors contributed as follows. Keerthana Deepti Karunakaran: conceptualization, methodology, software, formal analysis, investigation, writing–original draft, funding acquisition; Nancy D. Chiaravalloti: resources, writing–review and editing, supervision; Bharat B. Biswal: conceptualization, writing–original draft, funding acquisition, resources, supervision, project administration.

